# Multiple instance fine-mapping: predicting causal regulatory variants with a deep sequence model

**DOI:** 10.1101/2025.06.13.25329551

**Authors:** Alexander Rakowski, Christoph Lippert

## Abstract

Identifying causal genetic variants in a computational manner remains an open problem. Training end-to-end prediction models is not possible without large ground-truth datasets, while results of genome-wide association studies (GWAS) are entangled by linkage disequilibrium (LD), and gene expression datasets do not contain genetic variation at individual-level. Here, we propose Multiple Instance Fine-mapping (MIFM) – a multiple instance learning (MIL) objective to overcome the lack of strong labels by grouping putatively causal variants together based on their LD scores. Using MIFM, we trained a deep classifier on a dataset aggregating over 13, 000 GWAS to predict causal variants based on their underlying DNA sequences. We validated variants prioritized by MIFM by constructing polygenic risk scores which transferred better to different target ancestries. Furthermore, we demonstrated how MIFM can be used to disentangle effect sizes of highly-correlated variants to better fine-map GWAS results.

**Author summary:** Genome-wide association studies have identified tens of thousands genetic variants associated with traits or diseases. However, the majority of identified variants is only spuriously correlated with the phenotype of interest, having no causal effect on it. Instead, these variants are often inherited together with nearby biologically causal variants, thus creating the spurious associations. Fine-mapping, i.e., predicting which variants are causal, is crucial for downstream tasks, such as uncovering the biological mechanisms affecting the phenotype or robustly identifying individuals with high genetic risk of a disease. While most fine-mapping methods are based on the available association statistics or functional annotations of genetic regions, it should be possible to identify causal variants based on their neighboring DNA sequences. However, training a standard machine learning classifier for that task is obstructed by the scarcity of strong, ground-truth labels. Here, we proposed a method to train sequence models predicting variant causality using weakly-labeled data. We trained a model on a large set of associated variants, and demonstrated its utility by improving cross-ancestry predictions of genetic risk, or disentangling the effect sizes of highly correlated variants.

## 2 Introduction

Genome-wide association studies (GWAS) remain a powerful tool for identifying genetic variants associated with phenotypes or diseases, with recent studies detecting up to thousands of associations per trait. However, while one usually assumes a clear *genotype* → *phenotype* causal direction, only a small fraction of variants significant in a GWAS are expected to be truly causal [46]. Due to linkage disequilibrium (LD), single nucleotide polymorphisms (SNPs) in proximity to causal variants become associated with the trait and can even have lower p-values than the causal SNPs. Identifying the true causal variants is important for understanding the underlying mechanisms such as transcription factor (TF) binding and for making robust predictions in populations with different LD structures than the one where the GWAS was performed.

Without *in-vivo* experimental validation, one can employ computational *fine-mapping* methods, which aim to narrow down the set of putative causal variants from GWAS summary statistics. The simplest approach is to test the significance of all candidate SNPs in a joint model. While yielding unbiased estimates of the true effect sizes, it is not feasible in scenarios with large numbers of variants or strong LD. More advanced methods utilize the Bayesian framework to estimate credible sets of variants given prior knowledge on the distribution of effect sizes [22, 6, 51], optionally incorporating functional annotations as additional priors [26, 11, 55]. While powerful, the above methods require selecting hyperparameters, such as the assumed number of causal variants, rely on availability of functional annotations for the regions of interest, and are sensitive to LD patterns, yielding large credible sets for strongly correlated variants.

Another approach is to use machine learning (ML) prediction models to assign a score to each variant as a proxy of its likelihood of being causal. A common choice are deep neural networks (DNNs) trained to predict functional genomic annotations or gene expression values from DNA, with architectures typically based on a convolutional neural network (CNN) [59, 25, 3] or a transformer backbone [2]. The difference in predictions for a sequence with the reference allele and a sequence with the alternative allele is then taken as a measure of the potential causality of a variant. As opposed to the statistical methods, the ML-based approaches are independent of GWAS results or the LD structure. However, while they take DNA data at base pair level resolution as inputs, they are typically trained on reference genome data, which limits the SNP-level variability of DNA motifs to ones present at population-level. Furthermore, the performance of such models can be hindered by coarseness and noise of the labels [29, 8].

Here, we introduce Multiple Instance Fine-mapping (MIFM), a framework for training deep learning models to predict the causality of non-coding variants directly from the underlying DNA sequence, without the need for summary statistics or functional annotations at test time. We circumvent the lack of ground-truth labels at SNP resolution by formulating the training objective as a multiple instance learning (MIL) problem, where putatively causal variants in LD with each other are grouped together to form a single, weakly-labeled positive example, and fit the model on a dataset of more than 2 million associated variants from over 13, 000 studies. We demonstrate the robustness of MIFM-prioritized variants by creating polygenic scores (PGS) of 20 traits, which transferred better from European to non-European ancestries, compared to variants selected using existing fine-mapping methods. Furthermore, we show how MIFM can be used to detect additional signals in analyses of GWAS results by prioritizing variants for joint tests, even in strongly-correlated cases. Finally, we report the results of model analysis which revealed enrichment of regulatory elements, existing and putatively novel TF motifs, as well as context-dependent mechanisms. The corresponding code as well as the trained model used in our experiments are available at github.com/HealthML/multiple-instance-fine-mapping

## 3 Results

### 3.1 Overview of the method

We proposed Multiple Instance Fine-mapping (MIFM), a framework for training models prioritizing causal genetic variants with the multiple instance learning (MIL) paradigm, using GWAS associations as training data (Figure 1). Our goal was to obtain a classifier predicting the probability of a variant being causal, given the DNA sequence around it. To train such a model in the standard supervised manner one would need ground-truth labels for each individual *instance* (variant). However, variants discovered with GWAS typically contain a large number of false positives due to LD between SNPs. To circumvent the lack of ground-truth labels, we trained a model to classify *bags* of instances (LD blocks of variants), instead of individual SNPs. We constructed the training dataset by selecting significant variants from a large set of GWAS results and grouping SNPs in LD with each other into positive bags. Conversely, we created negative examples by selecting common variants from the human reference genome which were not significantly associated in any of the GWAS. During training, the model makes instance-level predictions for each SNP within an LD block, which are then pooled by selecting the highest score as the bag-level prediction. Once trained, the pooling operation is discarded, and the instance-level model can be used to make predictions for individual variants. See Section 5.1 for a detailed description of the method, and Section 5.2 regarding dataset construction.

**Figure 1:**
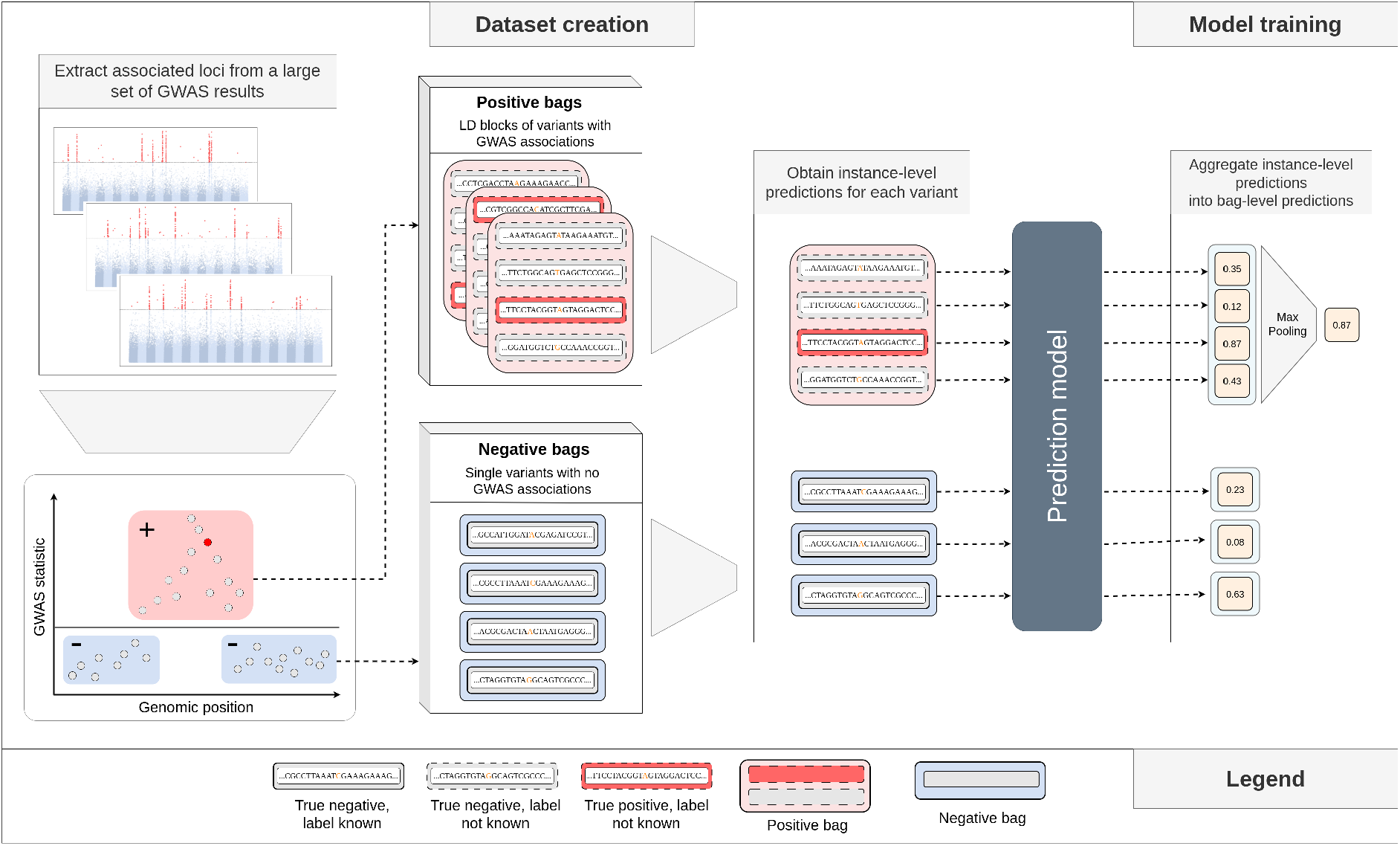
Overview of Multiple Instance Fine-mapping (MIFM) – a framework for training variant prioritization models. We frame the task of identifying causal variants as a multiple instance learning (MIL) problem, where loci of GWAS-associated SNPs are grouped to form positive “bags” for the MIL algorithm, to overcome the lack of instance-level (per-variant) labels. We assume that at each positive bag contains at least one causal variant, while negative bags contain none. **Dataset creation:** We construct the training dataset using a large set of GWAS results, by selecting SNPs significantly associated in any study (marked in red). Since we do not have variant-level labels, we treat whole LD-blocks of associated SNPs as positive bags. Conversely, we construct the set of negative examples (marked in blue) by selecting the remaining, non-significant variants, to form single-element negative bags. **Model training:** we train a prediction model, e.g., a deep neural network, to classify the MIL bags, based on the underlying DNA sequences of the variants. The model first makes separate predictions for each element in the bag, which are then aggregated using the *max* operator to yield a single bag-level prediction. After the model is trained, we can discard the *max* operation and predict the causality of single variants.

### 3.2 Polygenic risk scores created with MIFM transfer better to non-European ancestries

The predictive performance of PGS can decrease when applied to populations different from the one where the GWAS summary statistics were obtained from [33, 13, 32]. Due to varying LD patterns across ancestries, SNPs associated with the phenotype in the GWAS population might not be tagging the causal variants in the target population. As most studies are biased towards European populations [**?**, 47, 16], this can increase health disparities, e.g., by failing to identify individuals at risk in minority ancestries [34]. On the other hand, there is evidence for causal variants and their effect sizes being consistent across ancestries [54, 42, 23]. Thus, identifying causal variants should improve the cross-ancestry transferability of PGS.

We created PGS by prioritizing variants using MIFM and 9 baseline methods, and evaluated their performance on non-European ancestries (Section 5.3). Each method was evaluated for 20 traits and 5 ancestries, yielding a total of 100 scenarios per model. Within each scenario, we compared the performance of PGS created with MIFM and each baseline, and counted the total number of scenarios where MIFM would perform significantly better or worse than a baseline (Figure 2). Overall, MIFM performed better in 19% and worse in 5% of all scenarios. The net number of scenarios where MIFM performed better was positive regardless of the baseline, ranging from a net difference of 5% for ABF and Enformer, to 10% for Polyfun FINEMAP. The smallest improvements were obtained for the East Asian (EAS) ancestry (6% of scenarios better, 4% worse), while the largest improvements were for the African (AFR) ancestry (17% better, 2% worse) (Supplementary Figure 1). With respect to individual traits, MIFM performed the worst for breast cancer (4% of times better, 16% worse) and prostate cancer (11% worse), while the most consistent improvements were for serum urate levels (91% better) and coronary artery disease (CAD) (24% better) (Supplementary Figure 2).

**Figure 2:**
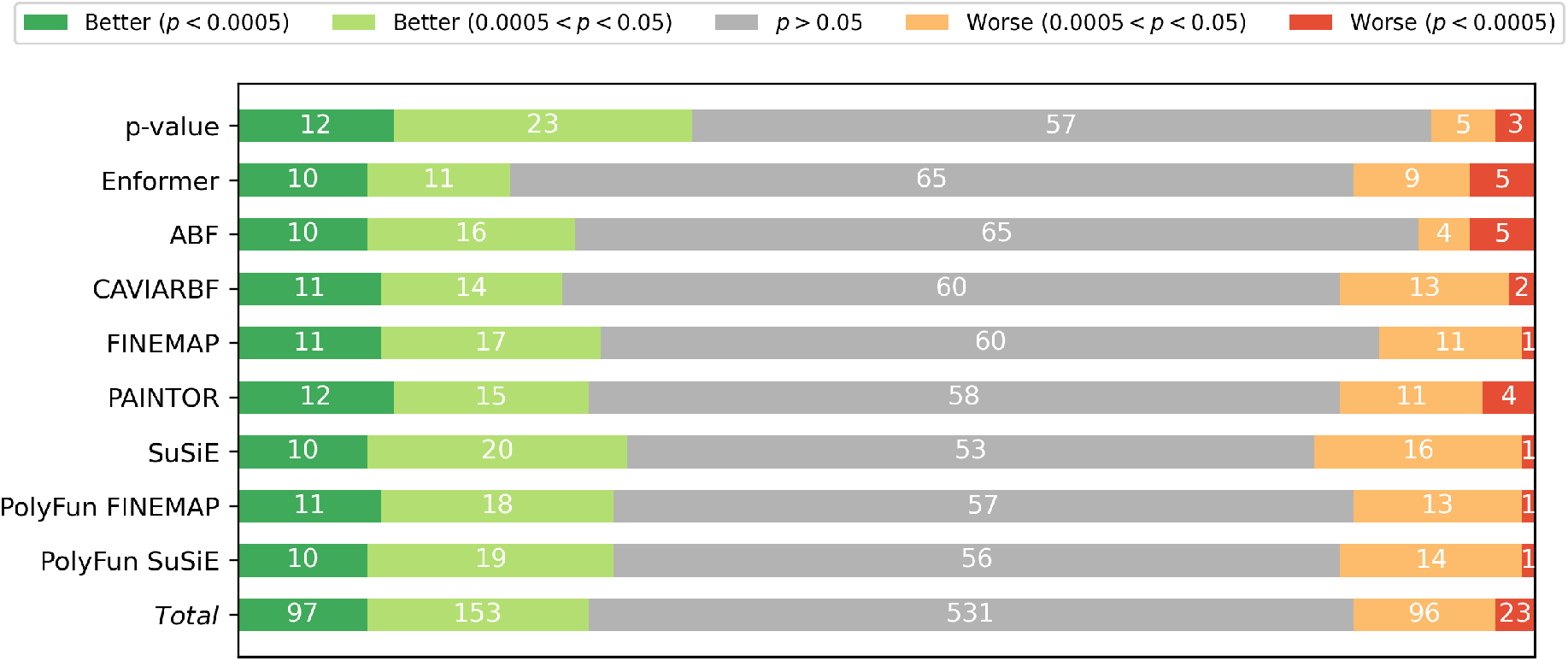
Performance comparison of PGS created with MIFM and 9 baseline methods on 5 non-European ancestries and 20 traits. We created PGS using results from 20 GWAS performed on European samples and evaluated them on 5 non-European samples, yielding 100 test scenarios per model. For each baseline, we counted the number of scenarios where MIFM would perform better than the baseline (in green), worse (in red), or not significantly different (in gray).

### 3.3 MIFM enables discovery of additional GWAS signals

Joint regression models of multiple variants can estimate the causal effect sizes instead of the marginal ones [56]. However, in the presence of a large number of highly correlated SNPs, large sample sizes are needed to disentangle the signals. Thus it is often infeasible to test all the putative variants jointly, especially for studies of modest sizes. We used MIFM to prioritize variants for conditional testing of highly-correlated (*R*^2^ ≥ 0.9) SNPs and compared the results with a naive approach of selecting all variants in high LD in 4 moderately sized GWAS in Table 1. In each GWAS, the joint regression of MIFM-prioritized SNPs yielded a larger number of significant variants, yielding 47 variants in total, compared to 32 variants from the baseline models. 2 out of the 47 variants had significantly different effect size estimates in the larger model, and might be false positives. One of the variants identified by the MIFM joint model was above the significance threshold in the GWAS and would otherwise be undetected by the marginal effect size estimates. We also counted secondary signals, i.e., cases where more than 1 SNP was significant in a joint model, where MIFM identified two cases more than the baseline models (Table 2).

**Table 1:** Number of variants identified by conditional analyses of GWAS results for selected phenotypes. We tested SNPs highly correlated with each lead SNP using a joint model of all variants in LD (2nd column) and of variants prioritized by p-values (3rd column) and by MIFM (4th column). In the 5th column we report the number of variants identified with MIFM whose effect size estimates were matching the estimates from the full model.

| Trait | # variants (baseline) | # variants (p-value) | # variants * (MIFM) | # variants ** (MIFM) |
| --- | --- | --- | --- | --- |
| Height | 9 | 7 | 13 | 13 |
| Red blood cell count | 9 | 10 | 12 | 12 |
| Systolic blood pressure | 0 | 0 | 1 | 1 |
| Heel bone mineral density | 14 | 15 | 21 | 19 |
| Total | 32 | 32 | 47 | 45 |

**Table 2:** Number of secondary signals identified by conditional analyses of GWAS results for selected phenotypes. We tested SNPs highly correlated with each lead SNP using a joint model of all variants in LD (2nd column) and of variants prioritized by p-values (3rd column) and by MIFM (4th column), and report the number of secondary signals, i.e., cases where more than one variant was significant in the joint model.

| Trait | # 2nd. signals<br>(baseline) | # 2nd. signals<br>(p-value) | # 2nd. signals<br>(MIFM) |
| --- | --- | --- | --- |
| Height | 0 | 0 | 1 |
| Red blood cell count | 1 | 1 | 0 |
| Systolic blood pressure | 0 | 0 | 0 |
| Heel bone mineral density | 2 | 1 | 4 |
| Total | 3 | 2 | 5 |

### 3.4 MIFM variants are enriched for enhancer, repressed, and silencer chromatin signatures

To characterize variants prioritized by MIFM, we analyzed whether they enrich for regulatory elements compared to all putatively causal variants from CAUSALdb2. We computed the enrichment for 20 GenoSTAN-defined states [57], which we divided into 6 groups: enhancers, promoters, repressed regions, transcriptional elongations, repressed-enhancer regions, and low-signal regions (Figure 3). MIFM-prioritized variants are enriched significantly for repressed regions (odds ratio (OR) from 1.02 for the lowest quantile to 1.13 for the highest quantile), repressed-enhancer regions (OR from 1.01 to 1.13), and the enhancer elements (OR from 1.01 to 1.10), with the highest enrichment for “strongly” defined enhancers subgroup (“Enh.6”, up to an OR of 1.16). Conversely, we observed a significant depletion of low-signal regions at higher model score quantiles (OR from 0.98 at the 0.7 quantile to 0.97 at the 0.9 quantile), and for transcription elongations (OR 0.99 for the 0.3 quantile to 0.98 for the 0.9 quantile), with the exceptions of “Gen5’.13” which was enriched for, with an OR up to 1.12. We did not observe significant deviations from 1 in the odds ratios for promoter regions. To understand why MIFM enriches for repressed regions, we further analyzed the repressed and repressed-enhancer variants prioritized by MIFM, and observed a significant enrichment for enhancer regions compared to all repressed and repressed-enhancer variants of CAUSALdb2 (up to 1.08 for repressed regions and up to 1.10 for repressed enhancer-like regions) (Supplementary Tables 1 and 2), indicating that repressed regions prioritized by MIFM are often active enhancers in other cell lines. Additionally, we analyzed repressed, repressed-enhancer, and enhancer regions in terms of silencers, and observed a significant enrichment in MIFM-prioritized repressed regions and in enhancer regions (up to 1.07 in both subsets), and no significant change in OR for repressed-enhancer regions (Supplementary Tables 3–5). The presence of silencers is associated with the H3K27me3 histone modification [9], which also characterizes the repressed GENOSTAN states, further suggesting that MIFM enriches for repressed regions with functional elements. Silencers can act as enhancers depending on the cellular context [19, 36, 12], and their enrichment in the enhancer subset can either indicate a preference of the model for such “dual” elements.

**Figure 3:**
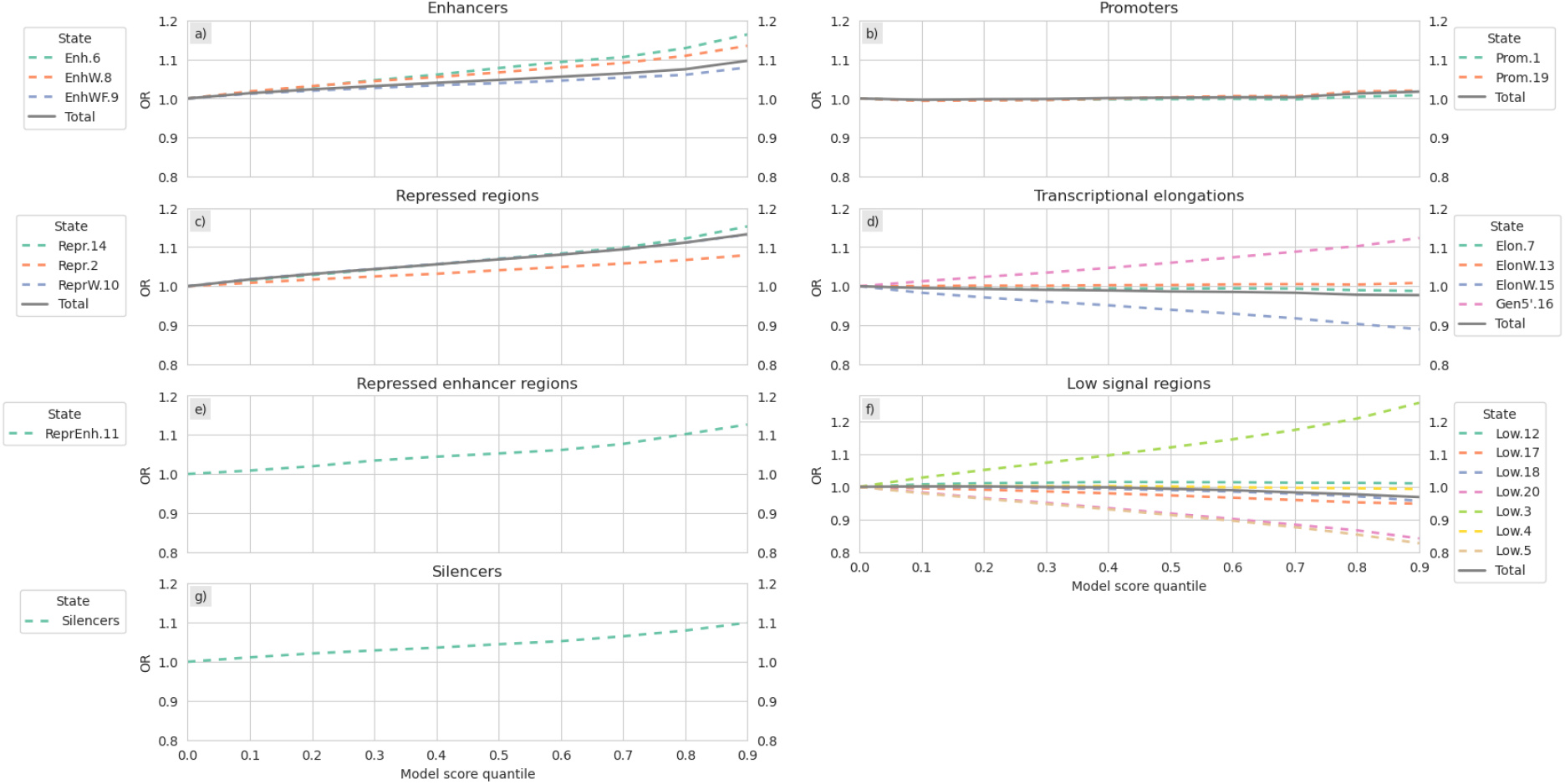
Enrichment analysis of regulatory elements in variants prioritized by MIFM. We plot the odds ratios (y-axis) of variants with MIFM scores above each quantile (x-axis) versus all variants in the CAUSALdb2 data. Each plot represents a different regulatory group based on GenoSTAN [57] chromatin state annotations (plots *a* to *f*) and silencerDB [58] validated and predicted silencer elements (plot *g*).

### 3.5 Syntax analysis of a MIFM trained model

We analyzed the trained MIFM model using Transcription-Factor Motif Discovery from Importance Scores (TF-MoDISco) and identified 161 unique DNA patterns in total, consisting of 67 patterns with positive attribution scores for models predictions and 94 patterns with negative attributions. The negative patterns had a smaller support in terms of corresponding seqlets (instances of similar patterns), with a median number of 798 seqlets compared to a median 1, 257 of seqlets for a positive pattern. In total, 60% of the positive patterns and 40% of the negative ones had a support of at least 1, 000 corresponding seqlet instances in the data. 20 positive and 49 negative patterns were significantly matched to at least one known human TF binding motif using TOMTOM [48]. TF motifs with the highest number of matched patterns are shown in Supplementary Table 6. Overall, we observed several motifs that were matched to both positive and negative patterns at the same time.

We further analyzed how predictions of MIFM are influenced by the positive and the negative patterns by performing in silico mutagenesis of sequences with patterns matching the binding motif of the ARID3A protein, a TF with reported interactions with other regulatory elements [18, 41, 43]. We modified sequences containing positive patterns upstream of the SNP position by replacing the positive pattern with the negative one, and vice versa, and compared MIFM predictions for the original and modified sequences (Figure 4). Adding negative patterns to sequences with a positive pattern shifted the distributions of scores towards lower values (from an average mean score of 0.54 to 0.48) and increased their spread (average standard deviation from 0.03 to 0.08).

**Figure 4:**
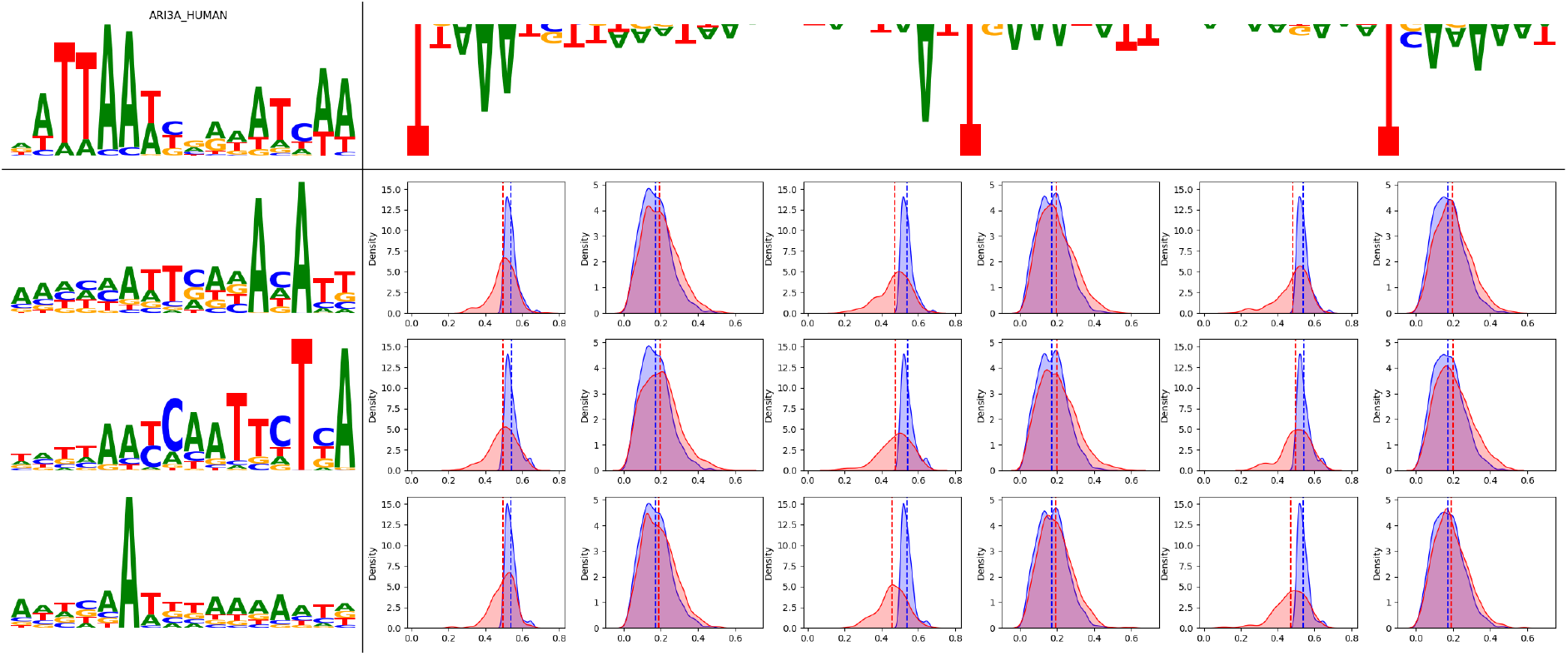
In silico mutagenesis of ARID3A-matching patterns and the corresponding MIFM predictions. We selected 3 positive patterns (rows) and 3 negative patterns (columns) matching the ARID3A motif which were identified by TF-MoDISco as influencing MIFM predictions. For each positive-negative pattern pair, we computed MIFM scores for sequences containing the positive pattern (odd subcolumns) and sequences with the negative pattern (even subcolumns) and plot the density functions of the scores in blue. We modified the sequences by adding the negative pattern to the positive sequences (odd subcolumns) and vice versa (even subcolumns), scored the modified sequences with MIFM, and plot the density functions of the modified scores in red. The vertical lines denote the means of MIFM scores of the original (blue) and modified sequences (red).

Adding positive patterns to “negative” sequences slightly shifted their scores towards higher values (average mean from 0.17 to 0.20, average standard deviation from 0.08 to 0.09). This suggests that the context of a variant is necessary, but not sufficient, for it to be predicted as causal by MIFM — one can “disable” the function of a variant by modifying its context, but it is not enough to modify the context to make a variant being predicted as causal.

## 4 Discussion

Identifying causal non-coding variants is typically done with fine-mapping methods which rely on summary statistics and population LD structure, without directly using the underlying DNA sequences. Alternatively, one can employ sequence models of gene expression, which, however, are trained on reference genome data and do not observe individual-level DNA variation. To this end, we proposed a problem formulation of fine-mapping using the multiple instance learning objective, where we predict the presence of GWAS associations within LD-blocks containing multiple variants. By using the underlying DNA sequences as input features we can exploit similarities in DNA patterns between causal variants, while by constructing the labels using GWAS summary statistics we indirectly incorporate the individual-level genetic variation which drives SNP associations.

Using this approach, we trained a DNN model which predicts the probability of a SNP being causal given its neighboring DNA sequence, allowing us to prioritize variants of interest. One of the motivations for identifying causal variants is to robustly predict genetic liability of a phenotype across different populations, especially those which are under-represented in GWAS. By evaluating MIFM prioritized variants across a range of traits and ancestries, we were able to increase the robustness of polygenic scores predictions compared to a wide range of baselines. Furthermore, we showed that utilizing sequence information can be useful for disentangling highly-correlated GWAS variants, a task otherwise statistically infeasible with typical sample sizes.

We note that our goal was to propose a framework for training variant-prioritization models, rather than developing a new DNN architecture. We employed a relatively lightweight model for our experiments (less than 25, 000 parameters), and while we did not observe improvements with more complex architectures, we did not conduct an exhaustive comparison of possible DNN models. Besides improving the predictive performance, a valuable extension would be to increase the interpretability of the model with interpretable-by-design architectures [37, 49]. We further note that as MIFM utilizes a database of GWAS results, it can be continuously fine-tuned whenever new summary statistics are available, each time further narrowing down the MIL objective.

We showed how one can utilize the vast amount of GWAS results available to train machine learning models for variant prioritization, overcoming the problem of inaccurate labels due to confounding from LD. Such models can complement traditional fine-mapping methods, being able to reduce the number of putative variants to be analyzed, even without the access to the corresponding test statistics. Finally, by introducing base-pair level variations in the training data, this paradigm can be used to increase the robustness of existing DNA sequence models.

## 5 Methods

### 5.1 Fine-mapping as a multiple instance learning problem

We begin by introducing the MIL paradigm for a binary classification task. We assume that the data consists of pairs of {**x**_*i,j*_, *y*_*i,j*_ }, where **x**_*i,j*_ ∈ ℝ^*d*^ are input features and *y*_*i,j*_ ∈ {0, 1} are binary labels, and the input instances are grouped into *bags* of examples **X**_*i*_ = {**x**_*i*,1_, … , **x**_*i,m*_}, where *m* can differ across **X**_*i*_. We use *i* to index along the bag-level (e.g., **X**_*i*_, *y*_*i*_) and *j* to index individual instances within a bag (e.g., **x**_*i,j*_, *y*_*i,j*_). At training time, we only have access to bag-level labels, which indicate whether at least one instance in a bag is positive, and are defined as:

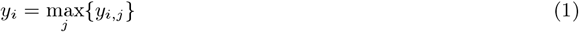

This can be interpreted as a form of weak labeling, since we do not know which particular instance(s) in a positive bag are positive, as opposed to strong, instance-level labels. Our goal is then to learn an instance-level classifier *f* : ℝ^*d*^ → [0, 1], given the bag-level data {**X**_*i*_, *y*_*i*_}. A common approach to train *f* is to define a bag-level classifier *F* :

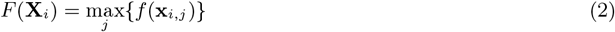

and train it to minimize the cross-entropy loss ℒ:

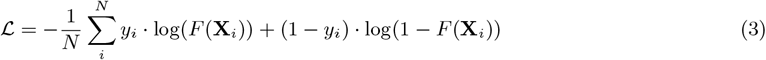

wrt. the bag-level examples {**X**_*i*_, *y*_*i*_}.

As GWAS estimate the marginal effect size of each SNP, the significantly associated variants typically comprise groups of SNPs in LD with each other, out of which only a small fraction is truly causal, and most SNPs are spuriously associated with the trait of interest through their correlations with causal variants. On the other hand, we assume that the causal signals are driven by DNA patterns around the variants, e.g., by TF binding motifs, and could be identified given enough data. Databases such as GWAS Catalog [10] or CAUSALdb [52] aggregate the results of thousands of GWAS, providing a large set of putatively causal variants of the human genome across a range of traits and populations. In order to use these data to infer about causal variants, we propose the following MIL scenario: let each **x**_*i,j*_ represent the DNA sequence centered at a SNP and *y*_*i,j*_ be unobserved ground-truth labels indicating causal variants. We construct positive bags 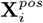 as independent LD-blocks of putatively causal variants, e.g., surpassing a given significance threshold in any study and grouped using a clumping procedure, while the negative ones are single-element bags 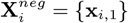 of variants without any significant associations. If at least one causal variant is present in each positive bag, this is a valid MIL objective. Assuming that the causal variants share DNA patterns between each other, we can successfully train a sequence model *f* , e.g., a neural network, to predict causal variants.

Formally, given *s* GWAS over *v* SNPs, let **S**_*i*_ ∈ ℝ^*v*^, *i* ∈ [*s*] be the summary statistics of the *i*-th GWAS and **S**_*i,j*_ be the p-value for the *k*-th variant in that study. Furthermore, let *b* be the total of independent LD-blocks, and *L*_1_, … , *L*_*b*_ be pair-wise disjoint sets of integers indicating which variants belong to which LD block. Given a significance threshold *T* , the positive bags are then defined as:

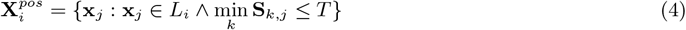

while the negative bags are defined as:

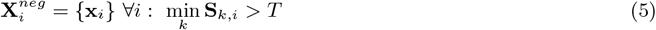

### 5.2 Dataset and model training

We constructed the training dataset using data from the CAUSALdb2 database [53], which aggregates 13, 709 GWAS summary statistics, resulting in over 2, 618, 834 putative causal variants from the GRCh37 human genome, which are grouped into 2, 772 independent LD-blocks. We further divided the blocks into primary and secondary signals, using labels provided by CAUSALdb2, yielding 4, 790 smaller blocks, and excluded blocks with fewer than 10 variants, to reduce the chance of a small number of false positive SNPs driving the training signal. We treated the resulting blocks as separate bag-level positive examples 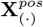. Since CAUSALdb2 only contains associated variants, we created the negative bags 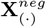 from common (MAF ≥ 0.01) GRCh37 variants which were not present in CAUSALdb2, excluding SNPs further than 128 base pairs away from any CAUSALdb2 variant, yielding a total of 1, 045, 506 training examples, each forming a separate single-element negative bag.

For model training, we employed a modified, smaller version of the Basenji2 CNN architecture [25], with 4 blocks in the first model stage, 2 blocks in the second stage, a final number of 64 filters, and a single output. We one-hot encoded the 512 base pair DNA sequences around each variant to serve as the inputs **x**_*i*_. As per Equation 2, for each block **X**_*i*_ = {**x**_*i*,1_, … , **x**_*i,m*_} the CNN makes a separate prediction *y*_*i,j*_ = *f* (**x**_*i,j*_) for each individual input **x**_*i,j*_ , and the maximum value across **x**_*i,j*_ ∈ **X**_*i*_ is taken as the bag-level prediction. We trained the model for 100 epochs using the Adam optimizer [27] with a learning rate of 10^−4^ and exponential decay of *γ* = 0.9 per epoch, with a batch size of 32 bags. For regularization, we used a dropout on the residual layer connections of the model with a rate of 0.5, and randomly shifted the input sequences by up to 8 base pairs. We used a modified DNA one-hot encoding by introducing a 6-th special token “V”, which we replaced the nucletotide value in the middle of the sentence, i.e., at the SNP position (*A* = [1, 0, 0, 0, 0], *C* = [0, 1, 0, 0, 0], *G* = [0, 0, 1, 0, 0], *T* = [0, 0, 0, 1, 0], *N* = [0, 0, 0, 0, 0], *V* = [0, 0, 0, 0, 1]). The use of this additional token allowed us to employ random shift augmentations by signaling the position of the variant of interest to the network, and thus potentially increasing its precision to the base pair level. Using the standard 5-token encoding (e.g., as in [2]) with random shifts would instead force the model to make the same predictions for any neighboring SNPs within the shift range of the variant of interest. To further increase robustness, we employed model ensembling [17] by repeating the training with 5 different random seeds, and training a student model [21] to predict the averaged output of the 5 models. We used the same network architecture for the student model with the exception of using the standard 5 token encoding and not using data augmentations. We implemented the models using the PyTorch [38] and PyTorch Lightning [14] software libraries, and trained them using a single NVIDIA A40 48GB GPU and 8 CPU cores per-model, with an average training time of 34 hours. We did not observe a benefit of using larger versions of the model, other network architectures (Enformer [2], BPNet [3], a pretrained MFD [40]), or differentiable alternatives to the *max* operator in the formulation of *F* in Equation 2 (log-sum-exponential [7], generalized mean [4], attention-based [24]). Finally, we note that since only “weak” labels are used for model fitting, the network can be further retrained, or fine-tuned, “at test time”, i.e., whenever one obtains results from a new GWAS, by updating the positive bags with the set of new putative variants.

### 5.3 Construction and evaluation of polygenic risk scores

We selected GWAS of 10 continuous and 10 binary traits from the CAUSALdb database [53] which were performed in populations of European ancestry and had matching traits in UK Biobank (UKB). For each study, we divided the corresponding variants into LD-blocks according to the CAUSALdb2 labels, treating the primary and secondary signals within a single block as separate blocks. We then constructed PGS by selecting the variant with the highest fine-mapping annotation score from each block. This resulted in 10 PGS per GWAS, using annotation scores from: MIFM, “raw” p-values, a pretrained Enformer model [2], and each of the 7 fine-mapping tools included in the CAUSALdb2 annotations: ABF [50], CAVIARBF [11], FINEMAP [6], PAINTOR [26], SuSiE [51], PolyFun FINEMAP [55, 6], and PolyFun SuSiE [55, 51]. For the raw p-values, we calculated the annotation scores as 1 minus the p-value of a variant. For Enformer, we calculated the annotation scores as the maximum differences in predictions for the alternative versus reference allele over all model outputs. We evaluated the scores on the African (AFR), Admixed American (AMR), Central/South Asian (CSA), East Asian (EAS) and Middle Eastern (MID) ancestry subsets of UKB, which we defined using the ancestry analysis functionality of pgs-calc [30], with a 1,000 Genomes LD reference panel [1]. This resulted in a total of 100 scenarios (20 traits × 5 ancestries). Within each scenario, we divided the samples into 5 folds and fitted 5 linear models of the PGS and covariates (age, sex, UKB assessment center, genotyping batch, and the first 10 genetic principal components). Each time we selected a different set of 4 folds for model training and the remaining fold for evaluation, and averaged the final outcome. For each of the 100 scenarios, we assessed the significance of the difference between the performances in terms of the *R*^2^ score (we used the McFadden pseudo-*R*^2^ [35] for binary traits) of MIFM and each baseline using a permutation test with 10^8^ permutations.

### 5.4 GWAS and conditional analyses

We performed GWAS of 4 traits — height, red blood cell count, systolic blood pressure, and heel bone mineral density — on a sample of *N* = 40, 000 unrelated individuals from UKB using the standard linear regression functionality of the BOLT-LMM software [31]. We filtered the SNPs with the following criteria: minor allele frequency (MAF)≥ 0.01, Hardy-Weinberg Equilibrium with a significance level of 10^−12^, and included imputed variants with an INFO score ≥ 0.01, which resulted in 9, 637, 426 SNPs in total. We transformed the phenotypes using the rank-based inverse normal transformation [5] and adjusted them for confounders using age, sex, the identifiers of the genotyping array and UKB assessment center, and the first 10 genetic principal components. For each trait, we constructed a set of independent loci using the clumping functionality of the PLINK software [39] with a significance threshold of 5 · 10^−8^ for the lead SNPs (variants with the lowest p-value per locus) and a threshold of 5 · 10^−6^ for secondary variants associated with the lead SNPs. Since we focus our analysis on disentangling the effect sizes of highly correlated variants, we used an *R*^2^ threshold of 0.9 and a physical distance threshold of 1, 000 kb. For each lead SNP and its secondary variants, we fitted three joint models of SNPs and confounders: a baseline model with all variants in the clump, and two models, which only included the lead SNP and variants filtered based on either their p-values of MIFM scores. For the filtered models we retained secondary variants with p-values below the 30-th percentile of p-values of all GWAS-associated SNPs, or above the 70-th percentile of scores for the MIFM model. For each locus we additionally fitted a full joint model of all variants on a second, independent sample, and tested for differences in effect size estimates of all significant variants from the previous step between the filtered, and full models. This was to exclude cases where a non-causal variant becomes significant as a “substitute”, due to the true causal variant beyond removed

### 5.5 Functional annotation of the training data

We utilized GenoSTAN [57] and silencerDB [58] data to annotate CAUSALdb2 variants in terms of their regulatory functions. For GenoSTAN data, we mapped all the variants from CAUSALdb2 into hg38 coordinates using the UCSC genome browser LiftOver tool [20] and annotated them with chromatin state annotations of 127 cell lines for the hg38 genome which we downloaded from https://www.cmm.in.tum.de/public/paper/GenoSTAN/. For each variant, we assigned to it the chromatin states which were repeated in at least 5 different cell lines. A single variant could thus have multiple assigned states due to heterogeneity of the cell line experiments. For example, it could be marked as an enhancer-like element in one experiment and marked as a repressed region in a different cell type. For silencerDB, we downloaded annotations for the GRCh37 genome from http://health.tsinghua.edu.cn/SilencerDB/download/Species/Homo_sapiens.bed, and marked all CAUSALdb2 variant within each silencerDB region as silencers. We performed enrichment analyses for each annotation class by computing the odds ratios of SNPs with the given label compared to a subset of variants passing a given MIFM score threshold, and computed the p-values for enrichment using the Fisher exact test [15].

### 5.6 Motif discovery

We used TF-MoDISco [3, 45] to identify motifs contributing to MIFM predictions. We computed the attribution scores for all training sequences using DeepLIFT [44] and ran TF-MoDISco with 200, 000 positive and 200, 000 negative seqlets (motif occurences). Finally, we matched the resulting patterns to known human TF binding models from HOCOMOCO 11 [28] using the TOMTOM [48]. For the in silico mutagenesis analysis of the contributions of positive and negative patterns, we selected sequences containing the 3 top positive and 3 top negative TF-MoDISco patterns matching the ARID3A binding motif. For each positive-negative pattern pair, we did the following:

1. We computed the offset with respect to the ARID3A motif for the positive and negative pattern. As we had access to the starting positions of the TF-MoDISco patterns for each example, we also computed the start position of ARID3A motif in them.
2. We obtained MIFM predictions for all positive and negative examples.
3. We selected the positions in the negative pattern where the probability of the top nucleotide exceeded 0.4.
4. For each positive example, we replaced the nucleotides at the positions matching the nucleotides selected from the negative pattern.
5. We computed MIFM predictions for the modified positive examples.
6. We repeated steps 3–5 by modifying the negative examples with the positive pattern.

## Data Availability

All data produced in the present study are available upon reasonable request to the authors

https://github.com/HealthML/multiple-instance-fine-mapping

